# Metabolomic and gut microbiome profiles across the spectrum of community-based COVID and non-COVID disease: A COVID-19 Biobank study

**DOI:** 10.1101/2022.08.07.22278510

**Authors:** Marc F. Österdahl, Ronan Whiston, Carole H. Sudre, Francesco Asnicar, Nathan J. Cheetham, Aitor Blanco Miguez, Vicky Bowyer, Michela Antonelli, Olivia Snell, Liane dos Santos Canas, Christina Hu, Jonathan Wolf, Cristina Menni, Michael Malim, Deborah Hart, Tim Spector, Sarah Berry, Nicola Segata, Katie Doores, Sebastien Ourselin, Emma L Duncan, Claire J Steves

## Abstract

Whilst many with SARS-CoV-2 infection have mild disease, managed in the community, individuals with cardiovascular risk factors experienced often more severe acute disease, requiring hospitalisation. Increasing concern has also developed over long symptom duration in many individuals, including the majority who managed acutely in the community. Risk factors for long symptom duration, including biological variables, are still poorly defined.

We examine post-illness metabolomic and gut-microbiome profiles, in community-dwelling participants with SARS-CoV-2, ranging from asymptomatic illness to Post-COVID Syndrome, and participants with prolonged non-COVID-19 illnesses. We also assess a pre-established metabolomic biomarker score for its association with illness duration.

We found an atherogenic-dyslipidaemic metabolic profile, and greater biomarker scores, associated with longer illness, both in individuals with and without SARS-CoV-2 infection. We found no association between illness duration and gut microbiome in convalescence.

Findings highlight the potential role of cardiometabolic dysfunction to the experience of long illness duration, including after COVID-19.

## Introduction

The devastation caused by the COVID-19 pandemic is unprecedented in recent memory, with >6.3 million deaths and 543 million cases worldwide in just over two years^1^. SARS-CoV-2 infection can cause a wide spectrum of illness, even in individuals who do not require acute hospital management: many individuals are asymptomatic (35.1% to 40.5% in meta-analyses^2,3^) while others report prolonged symptom duration (2.3% to 37.7%^4^). Post-COVID syndrome (PCS84)^5 6^ is defined by the National Institute for Health and Care Excellence (NICE) as: “signs and symptoms that develop during or after an infection consistent with COVID⍰19, continue for more than 12 weeks (84 days) and are not explained by an alternative diagnosis”^5^. A further category Ongoing Symptomatic COVID (OSC28) is defined by NICE as symptoms for between 4 weeks and 12 weeks (28-83 days). Together, OSC28 and PCS84 are known colloquially as ‘Long COVID’. The strongest predictors of PCS84 are age (with those aged 35-69 years having highest risk), female sex, and greater severity of acute infection^7,8^ Whilst vaccination against SARS-CoV-2 reduces the risk and duration of PCS84^9–11^, prolonged post-infection symptomatology remains common. The United Kingdom Office for National Statistics report 2 million affected individuals in the United Kingdom by 01 May 2022, and 71% of individuals report that this affects normal daily activities^12^. The understanding of the pathophysiology and risk factors for these differing phenotypes – from asymptomatic infection to prolonged illness - is still evolving.

Early in the pandemic, it was noted that individuals with cardiovascular disease were at greater risk of severe illness^13,14^. Metabolomic profiles, particularly lipidomics, can identify risk of cardiovascular disease, with known associations of particular profiles with cardiovascular disease^15^ and Type 2 diabetes^16,17^. Such pre-pandemic lipidomics profiles have been associated with risk of hospitalisation for both COVID-19, and pneumonia caused by other pathogens, enabling the generation of an Infectious Diseases risk score (ID-score) for hospitalisation due to COVID-19^18^. Disturbances to the same group of metabolites have also been observed in samples from hospitalised individuals when acutely unwell with COVID-19^19 20^. What is less clear is whether such metabolomic profiles are associated with disease duration, and/or Long-COVID, with the published studies focusing on specific metabolites in hospitalised individuals^21^.

However, most cases of COVID-19 are managed in the community rather than hospital, and we aimed to assess whether metabolomic profiles differed in community-dwelling individuals with different symptom durations, comparing samples from asymptomatic individuals, those with short duration, OSC28 and PCS84. In parallel, we analysed samples taken from individuals with acute illness of varying duration prompting a test which was negative, and later antibodies also indicated no evidence of prior SARS-CoV-2 infection. We examined metabolites individually and assessed the previously published ID-score.

Metabolism has been related to the gut microbiome composition, with reports that the gut microbiome may separate individuals with PCS84 from healthy controls^22,23^. Therefore, we further explored whether stool microbiome composition, taken after acute illness, was different between individuals with disease of different symptom duration with and without previous, confirmed SARS-CoV-2 infection. Finally, we tested whether there was any relationship between metabolomic profiles and gut metagenomic composition.

## Materials and Methods

### Cohort description

Study participants were volunteers from the COVID Symptom Study Biobank (CSSB, approved by Yorkshire & Humber NHS Research Ethics Committee Ref: 20/YH/0298). Individuals were recruited to the CSSB via the ZOE COVID Study^24^ (ZCS) using a smartphone-based app developed by Zoe Ltd, King’s College London, the Massachusetts General Hospital, Lund University, and Uppsala University, launched in the UK on 24 March 2020 (approved by the Kings College London Ethics Committee LRS-19/20-18210). Via the app, participants self-report demographic information, symptoms potentially indicative of COVID-19 disease (both closed/polar questions, and free text), any SARS-CoV-2 testing, and SARS-CoV-2 and influenza vaccinations. Participants can be invited via email to participate in other studies, according to eligibility.

In October 2020, prior to UK vaccination roll-out, 15,564 adult participants from the ZCS were invited to join the CSSB. Invited participants had: (a) a self-reported SARS-CoV-2 test result: RT-PCR at the start of illness, or a subsequent antibody test, whether positive or negative; and (b) logged at least once every 14 days since start of illness, or since the start of logging if asymptomatic.

Initially, individuals were recruited in four groups based on understanding of Long COVID at that time, and prior to definitions being published: (1) Asymptomatic, with confirmed infection; (2) Short illness (≤14 days) after confirmed infection; (3) Long illness (≥28 days)^5^ after confirmed infection; and (4) Long symptom duration (≥28 days) but with a negative test for SARS-CoV-2 infection (Table 1). Invited individuals were matched across these four groups by Euclidian distance for age, sex and BMI^9^. Participants were invited by email, and consented separately into the CSSB. Participants were sent home sampling kits in November 2020 via post, and returned capillary blood samples for metabolomic analysis. This also enabled antibody testing using an ELISA method^25^ to confirm prior infection status of all participants, the current standard for retrospective ascertainment^26^. A subgroup also consented to send in stool samples for analysis of their gut microbiome.

**Table 1:** Table describing definitions used to group individuals based on their swab RT-PCR/antibody test status and duration of symptoms, as logged in ZOE COVID Symptom Study app. Intermediate groups show in S Table 1.

| Group | For Swab test | For Antibody Test - If no appropriate swab available |
| --- | --- | --- |
| Asymptomatic COVID-19 | Positive swab with no symptoms around test (14 days before to 7 days after inclusive), with logging at least every 7 days | Positive antibodies with no symptoms before test including in "past symptoms", with logging at least every 7 days |
| Acute COVID-19 illness (ACI) | Positive swab test with swab taken up to 7 days prior, or 14 days after onset of symptoms. | Positive antibody test, with symptomatic symptoms lasting 7 days or less, starting >14 days before testing |
| Ongoing symptomatic COVID-19 (OSC28) | Positive swab, around onset of symptoms (as above). Symptoms lasting over 28 days up to 84 days. | Positive antibody test, with symptoms lasting over 28 up to 84 days, starting >14 days before testing |
| Post COVID-19 syndrome (PCS84) | Positive swab test with symptoms lasting ≥84 days | Positive antibody test, with symptoms lasting ≥84 days starting >14 days before testing. |
| Negative Non-COVID-19 illness 28-83 days (NC28) | Negative swab test in first 2 weeks of symptoms, and symptoms lasting 28 to 83 days inclusive. No other positive test during logging, including a negative antibody test at enrolment | Negative antibody test >14 days after onset of symptoms, with symptoms lasting 28 to 83 days inclusive |
| Negative Non-COVID-19 illness ≥84 days (NC84) | Negative swab test in first 2 weeks of symptoms, and symptoms lasting ≥ 84-days. No other positive test during logging, including a negative antibody test at enrolment. | Negative antibody test >14 days after onset of symptoms, with symptoms lasting ≥ 84 days |

Study participants were subsequently aligned using symptoms ascertained up to sample collection date, and the permissible gap in logging was further tightened to 7 days to increase accuracy of classification. Long-COVID groups were reshaped to match definitions published in November 2020^5^(see Table 1) corresponding to OCS (28-83 days, OSC28) and Post-COVID-19 Syndrome (>84 days, PCS84). The same duration parameters were applied to those reporting symptoms with the same timeframe parameters around a negative test for SARS-CoV-2, who were presumed to have a non-COVID-19 illness. This yielded six groups for comparison - four SARS-CoV-2 positive groups: Asymptomatic, Acute COVID-19 (≤7 days), OSC28, PCS84; and two SARS-CoV-2 negative groups: Non-COVID-19 illness 28-83 days (NC28), Non-COVID-19 illness ≥84 days (NC84).

To check that groupings assigned by the recruitment algorithm were clinically accurate, symptom logging maps were scrutinised in a subsample (n=115) by two researchers (MFÖ, CJS), independently and blind to algorithmic phenotype classification, before analysis. Final categories are detailed in Table 1.

Due to changes in logging stringency criteria, some participants also fell into additional, shorter categories of illness duration, detailed in Supplementary Table 1. These additional phenotypes have been reported in supplementary data tables, but not included in primary analysis as they were not recruited for this purpose, and their classification is less certain.

### Metabolomics

Capillary blood samples were obtained, between November 2020 and January 2021, when the majority of participants had recovered. Samples were returned in plasma collection tubes with initial processing of 20µL used for serology with the remainder frozen. Samples were processed in March/April 2021 by Nightingale Health Oyj (Helsinki, Finland) using high-throughput nuclear magnetic resonance metabolomics, measuring 249 metabolites including lipids, lipoprotein subclasses with lipid concentrations within fourteen subclasses, lipoprotein size, fatty acid composition, and various low-molecular weight metabolites including amino acids, ketone bodies and glycolysis metabolites^27^. Of these, 37 are certified for clinical diagnostic use and formed the focus of our analysis (referred to herein as “Clinically Validated”)^28^. Quality control was performed and reported by Nightingale Health. Due to postal transit time, glucose, lactate, and pyruvate could not be assessed and have been excluded from analyses, and creatinine was unavailable. There were no concerns raised with other biomarkers. Metabolites measured using this panel have been associated with the risk of hospitalisation for COVID-19 in the UK Biobank previously ^18^, including 25 of the clinically validated biomarkers used in an Infectious Diseases risk prediction score (ID Score) derived using Lasso regression^18^.

### Gut Microbiome

#### Sample Collection and Faecal Sample Processing

Two faecal samples per individual were collected and returned by post: faecal material from both collection tubes were homogenised in a Stomacher® bag, aliquoted out and stored at -80 degrees Celsius. The first 301 samples that would maintain a balance for BMI, age, and sex, were selected to undertake a pilot investigation of microbiome differences.

#### DNA extraction and sequencing

Genomic DNA (gDNA) was isolated from 1g faecal sample, using a modified protocol of the MagMax Core Nucleic Acid Purification Kit and MagMax Core Mechanical Lysis Module^29^. Libraries were prepared using the Illumina DNA Prep (Illumina Inc., San Diego, CA, USA) following the manufacturer’s protocol. Libraries were sequenced (2⍰×⍰150⍰bp reads) using the S4 flow cell on the Illumina NovaSeq 6000 system.

#### Metagenome quality control and pre-processing

All metagenomes were quality controlled using the pre-processing pipeline (available at https://github.com/SegataLab/preprocessing). Briefly, pre-processing consisted of three main steps: (i) read-level quality control, (ii) removal of host sequence contaminants, and (iii) splitting and sorting of cleaned reads. Read-level quality control removes low-quality reads (quality score <Q20), fragmented short reads (<75⍰bp), and reads with ambiguous nucleotides (>2 Ns), using trim-galore (https://www.bioinformatics.babraham.ac.uk/projects/trim_galore/). Host sequences contaminant DNA were identified using Bowtie 2^30^ with the “--sensitive-local” parameter to remove both the phiX 174 Illumina spike-in and human-associated reads. Splitting and sorting allowed for creation of standard forward, reverse, and unpaired reads output files for each metagenome.

#### Taxonomic and functional profiling

The metagenomic analysis was performed using the bioBakery 3 suite of tools^31^. Taxonomic profiling and estimation of species’ relative abundances were performed with MetaPhlAn 3 (v. 3.0.7 with “--stat_q 0.1” parameter)^31,32^. MetaPhlAn 3 taxonomic profiles were used to compute three alpha diversity measures: (i) the number of species with positive relative abundance in the microbiome (‘Richness’), (ii) the Shannon diversity index, independent of richness, which measures how evenly microbes are distributed (‘Shannon’)^33^, and (iii) the Simpson diversity index, which accounts for the proportion of species in a sample (‘Simpson’)^34^. Similarly, species-level relative abundances were used to estimate microbiome dissimilarity between participants (beta diversity) using the Bray-Curtis dissimilarity metric, which accounts for the shared fraction of the microbiome between two individuals and their relative abundance values^35^. Functional potential profiling of metagenomes was performed with HUMAnN 3 (v. 3.0.0.alpha.3 and UniRef database release 2019_01)^31,36^ that produced pathway profiles and gene family abundances. We assessed beta diversity by computing a Principal Coordinates Analysis (PCoA) based on the pairwise Bray-Curtis dissimilarity metric.

#### Additional Covariates

BMI was derived from self-reported weight and height. Other self-reported information (obtained from ZCS app-reported data) included smoking; and co-morbid illness (‘heart disease’ (otherwise unspecified), ‘diabetes’ (and type of diabetes), ‘lung disease’ (including asthma), hay fever, eczema, ‘kidney disease’ (otherwise unspecified) and current cancer (type, and cancer treatment, unspecified). Address data was linked to the UK Index of Multiple Deprivation (IMD), with the IMD rank decile used as a categorical variable measuring local area deprivation^37–40^. Frailty was assessed using the Prisma-7 scale, with a score >2 indicating frailty^41^.

A subset of participants had participated in a dietary assessment during the COVID-19 pandemic, also recruited through the ZCS (published previously^42,43^). This included detailed information on vitamin supplementation (including omega-3 oils), physical activity, alcohol consumption, dietary habits and a food frequency questionnaire. These data were used to derive a diet quality score^42^, and a plant-based diet index^42^, analysed as continuous variables. Both have previously been associated with cardiovascular disease^44^, Type 2 Diabetes^45^, a lower risk of COVID-19 illness, and a lower risk of hospitalisation for COVID-19 during the early waves of the pandemic^42^.

#### Statistical analysis

The statistical analyses were performed using R software (v. 4.0.5) and Stata (v.17, StataCorp).

Baseline characteristics were described by frequency and percentages. Descriptive data on those invited and those enrolled, are presented in Supplementary Table 2 + 3. Metabolites were all log-transformed and standardised (mean 0, standard deviation 1) as per protocol^18^. To account for 0 values, prior to log transformation, a pseudo-count of 1 was added to all values.

Initial analysis examined association between COVID-19 phenotypes and each metabolite individually, and the ID score^18^, using multinomial logistic regression (all adjusted for age, sex, and BMI), with the Asymptomatic group as the reference category of the outcome variable. The Benjamini-Hochberg False discovery rate method was used to correct p-values for multiple testing^46^.

To assess potential confounders, we performed eight sensitivity analyses additionally adjusting for: (1) self-reported co-morbidities (cardiovascular disease and diabetes), (2) Frailty, (3) IMD rank decile, (4) lifestyle variables (smoking status, frequency of alcohol consumption, frequency of physical activity), (5) self-reported use of any health supplement, (6) self-reported use of Omega-3 containing supplements, (7) Diet quality score, and (8) Healthy plant-based diet index (hPDI).

Differences between the alpha diversity distributions of the groups were assessed using the Wilcoxon rank-sum test within the ‘RClimMAWGEN’ package (p-value≤0.05 considered significant). With a sample size of 300 individuals, we have 79% power at 0.05 significance level, assuming a low effect size of 0.20. PERMANOVA from the ‘adonis2’ function of the ‘vegan’ package, was used to test for differences between groups based on the beta diversity computed from the PCoA of the pairwise Bray-Curtis dissimilarities. A generalized linear model was built using the ‘glm’ function of the ‘stats’ package, controlling for confounding factors, including age, sex,and BMI. Only species with minimum 20% prevalence were used in this statistical analysis^47^. P-values were corrected using the Benjamini-Hochberg method^46^.

Spearman correlation analyses were conducted to associate microbiome profiles of 301 individuals with their metabolome profiles, adjusting for confounding factors (age, sex, BMI). Correlation analyses were conducted using R version 3.6.0. The package ‘corrplot_0.90’ was used to compute the variance and the covariance or correlation, ‘pheatmap_1.0.12’ and ‘cor.mtest’ were used to visualise the heatmap, calculate associated p values, and ‘p.adjust’ was used to perform Benjamini-Hochberg multiple testing correction^46^.

## Results

### Baseline Characteristics of Cohort

Of 15 564 individuals invited to the CSSB, 5694 (36.6%) consented and were enrolled. Of these, 4787/5694 individuals (84.1%) returned samples suitable for metabolomic analysis. Participant mean age was 52.5 years (SD 11.8), 78.7% were female, and 94.8% identified as White British, (Table 2a and Supplementary Table 2).

**Table 2a:** Baseline characteristics of each illness category. n: number; sd: standard deviation; IQR: Inter-quartile range.

|  | Asymptomatic |  | Acute COVID-19<br>(≤ 7 days) |  | Ongoing Symptomatic<br>COVID-19 (28-83 days) |  | Post COVID-19<br>Syndrome (≥84 days) |  | Non-COVID-19<br>illness (28-83 days) |  | Non-COVID-19 illness<br>(≥84 days) |  |
| --- | --- | --- | --- | --- | --- | --- | --- | --- | --- | --- | --- | --- |
|  | n | Col % | n | Col % | n | Col % | n | Col % | n | Col % | n | Col % |
| **Total** | 307 |  | 1147 |  | 652 |  | 180 |  | 239 |  | 48 |  |
| **Age (mean, sd)** | 58.1 | (10.2) | 53.2 | (12.0) | 53.1 | (11.2) | 53.6 | (11.5) | 53.7 | (11.0) | 58.1 | (9.2) |
| **Age Groups** |  |  |  |  |  |  |  |  |  |  |  |  |
| **<30** | 5 | 1.6% | 41 | 3.6% | 19 | 2.9% | 7 | 3.9% | 5 | 2.1% | 0 | 0.0% |
| **30-39** | 9 | 2.9% | 130 | 11.3% | 65 | 10.0% | 16 | 8.9% | 26 | 10.9% | 1 | 2.1% |
| **40-49** | 47 | 15.3% | 219 | 19.1% | 133 | 20.4% | 36 | 20.0% | 42 | 17.6% | 7 | 14.6% |
| **50-59** | 92 | 30.0% | 401 | 35.0% | 247 | 37.9% | 67 | 37.2% | 88 | 36.8% | 18 | 37.5% |
| **60-69** | 122 | 39.7% | 259 | 22.6% | 151 | 23.2% | 41 | 22.8% | 63 | 26.4% | 17 | 35.4% |
| **70+** | 32 | 10.4% | 95 | 8.3% | 37 | 5.7% | 13 | 7.2% | 14 | 5.9% | 5 | 10.4% |
| **Sex** |  |  |  |  |  |  |  |  |  |  |  |  |
| **Male** | 68 | 22.1% | 251 | 21.9% | 144 | 22.1% | 35 | 19.4% | 39 | 16.3% | 12 | 25.0% |
| **Female** | 238 | 77.5% | 892 | 77.8% | 505 | 77.5% | 141 | 78.3% | 200 | 83.7% | 36 | 75.0% |
| **Other/PFNTS** | 1 | 0.3% | 4 | 0.3% | 3 | 0.5% | 4 | 2.2% | 0 | 0.0% | 0 | 0.0% |
| **BMI (median, IQR)** | 25.3 | (25.6-28.4) | 25.2 | (22.7-28.6) | 25.5 | (22.9-29.4) | 26.3 | (23.4-31.9) | 25.7 | (22.7-30.3) | 26.2 | (22.5-29.7) |
| **<18.5** | 1 | 0.3% | 10 | 0.9% | 5 | 0.8% | 5 | 2.8% | 6 | 2.5% | 1 | 2.1% |
| **18.5-24.9** | 141 | 45.9% | 547 | 47.7% | 287 | 44.0% | 59 | 32.8% | 103 | 43.1% | 18 | 37.5% |
| **25-29.9** | 111 | 36.2% | 369 | 32.2% | 210 | 32.2% | 62 | 34.4% | 65 | 27.2% | 18 | 37.5% |
| **30.0-34.9** | 36 | 11.7% | 139 | 12.1% | 95 | 14.6% | 33 | 18.3% | 42 | 17.6% | 7 | 14.6% |
| **35+** | 13 | 4.2% | 75 | 6.5% | 49 | 7.5% | 21 | 11.7% | 21 | 8.8% | 4 | 8.3% |
| **Missing** | 5 | 1.6% | 7 | 0.6% | 6 | 0.9% | 0 | 0.0% | 2 | 0.8% | 0 | 0.0% |
| **Test (n, % confirmed by PCR)** | 15 | 4.9% | 567 | 49.4% | 315 | 48.3% | 84 | 46.7% | 163 | 68.2% | 28 | 58.3% |

**Table 2b:** Microbiome cohort demographics (Age, Sex, BMI) per COVID group.

|  | Asymptomatic | Acute COVID-19 | Ongoing Symptomatic | Post COVID-19 | Non-COVID-19 |
| --- | --- | --- | --- | --- | --- |

**Table 2b:** Microbiome cohort demographics (Age, Sex, BMI) per COVID group.
|  |  |  | (≤ 7 days) |  | COVID-19 (28-83 days) |  | Syndrome (≥84 days) |  | illness (≥28 days) |  |
| --- | --- | --- | --- | --- | --- | --- | --- | --- | --- | --- |
|  | n |  | n |  | n |  | n |  | n |  |
| Total | 35 |  | 109 |  | 52 |  | 20 |  | 85 |  |
| Age (Median, IQR) | 57 | (4.75) | 57 | (4.86) | 57 | (5.67) | 53 | (3.63) | 55 | (5.42) |
| Sex |  |  |  |  |  |  |  |  |  |  |
| Male | 5 | 14% | 28 | 26% | 14 | 27% | 8 | 40% | 15 | 18% |
| Female | 30 | 86% | 81 | 74% | 38 | 73% | 12 | 60% | 70 | 82% |
| BMI (Median, IQR) | 24.3 | (4.75) | 24.9 | (4.86) | 24.5 | (5.67) | 26.5 | (3.63) | 24.0 | (5.42) |

### Metabolomic Analysis

In total, 3718/4787 (77.7%) participants had adequate logging and metabolomic data s, of whom 2561/3178 (80.6%) fell into the defined phenotype groups (see Table 1). Baseline characteristics of the groups were broadly similar to the population from which they were recruited and to each other, although the final asymptomatic group was slightly older than the average for the cohort (mean 58.1 years (SD 10.2) vs. 52.7 (SD 11.7) p<0.001) (Table 2a, Supplementary Table 2 + 3).

Regression analysis, adjusting for age, sex, and BMI, showed 90 of 249 (36%) metabolites differed in participants with OSC28 (28-83 days of COVID-19 symptoms) compared to Asymptomatic SARS-CoV-2-positive individuals (Figure 1, Supplementary Table 4). 39 of these 90 (43%) also differed in participants with NC28 (Non-COVID-19 illness 28-83 days) compared to Asymptomatic SARS-CoV-2 infection, with the same direction of effect (Figure 1, Supplementary Table 4).

**Figure 1:**
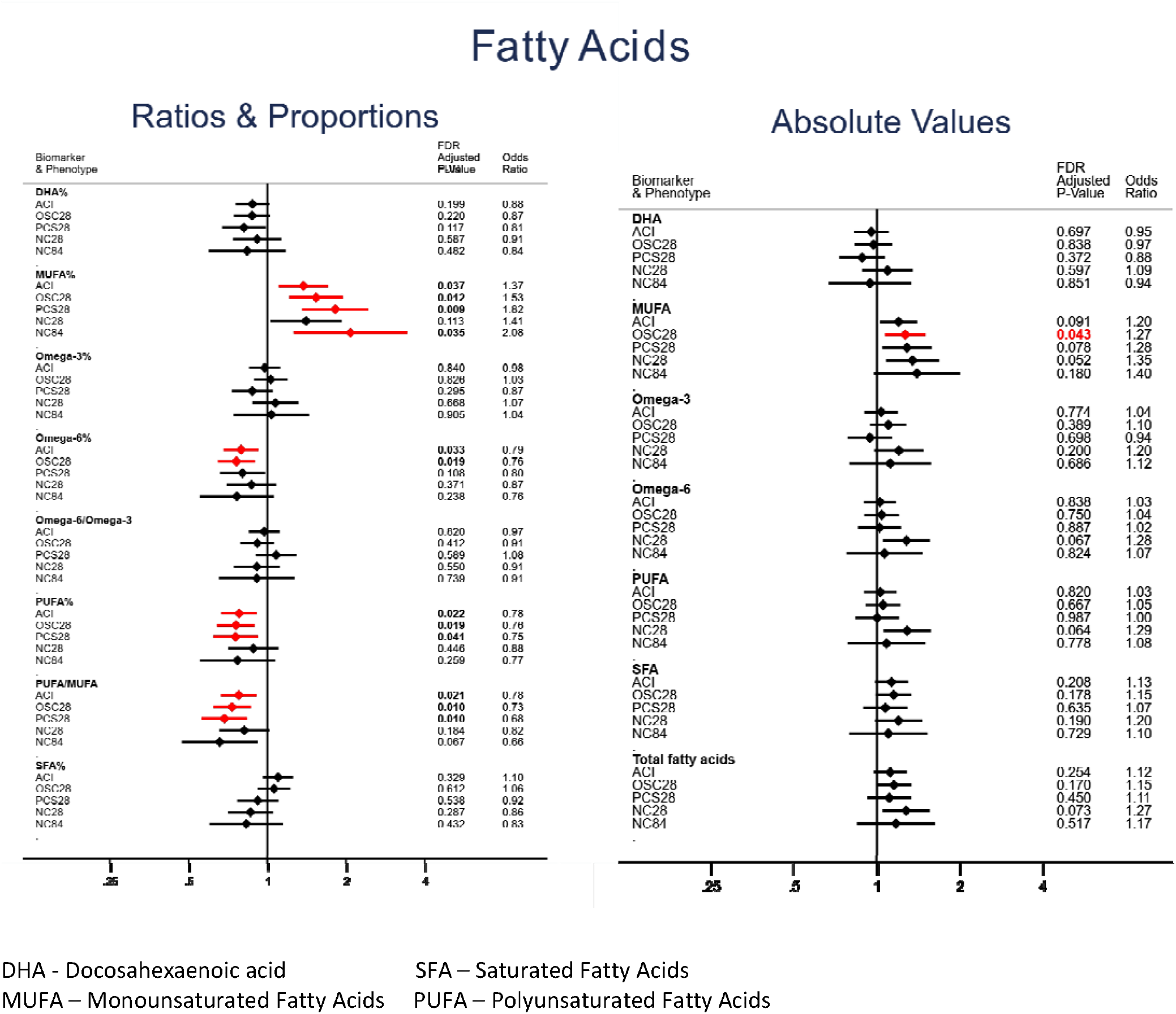
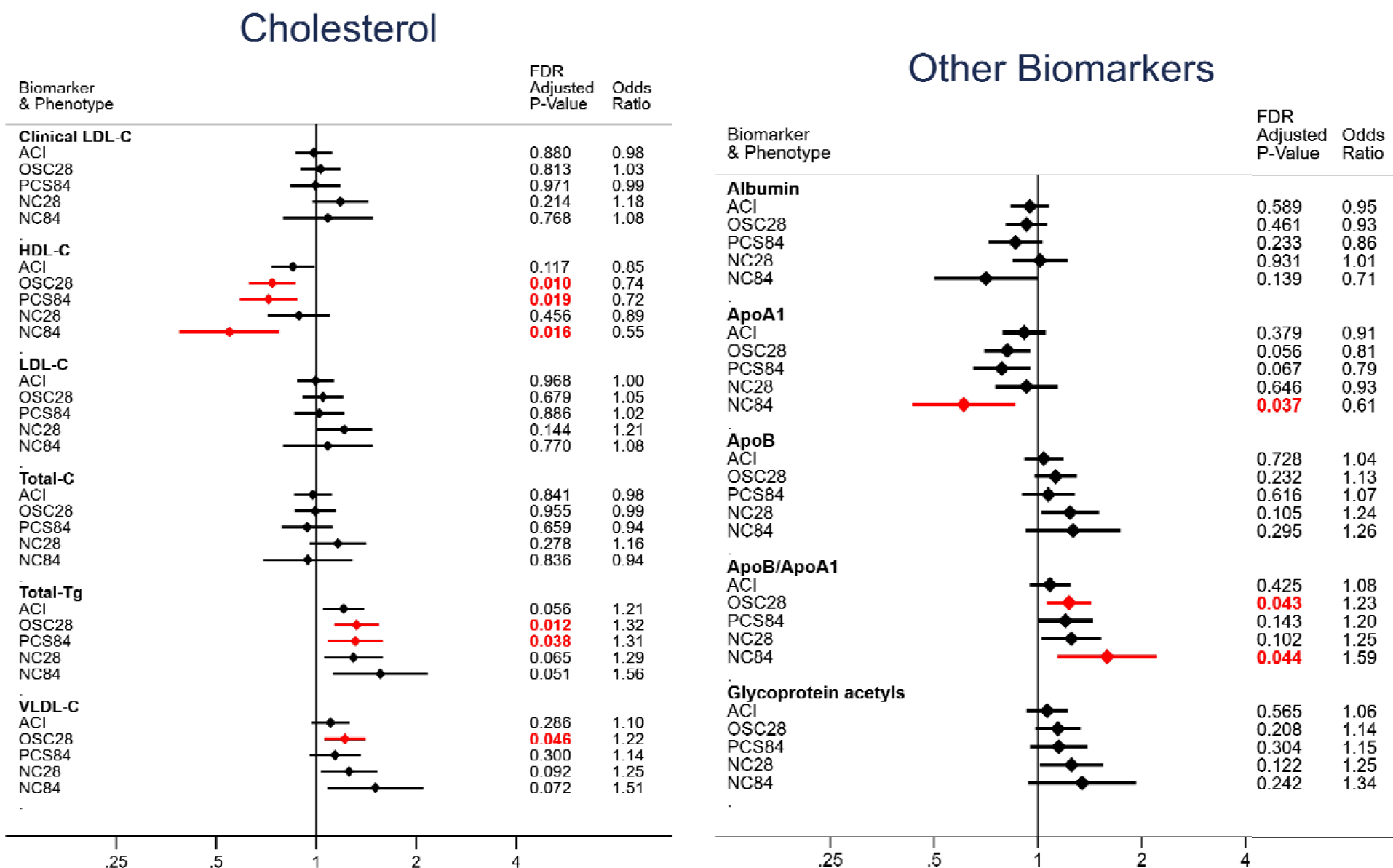
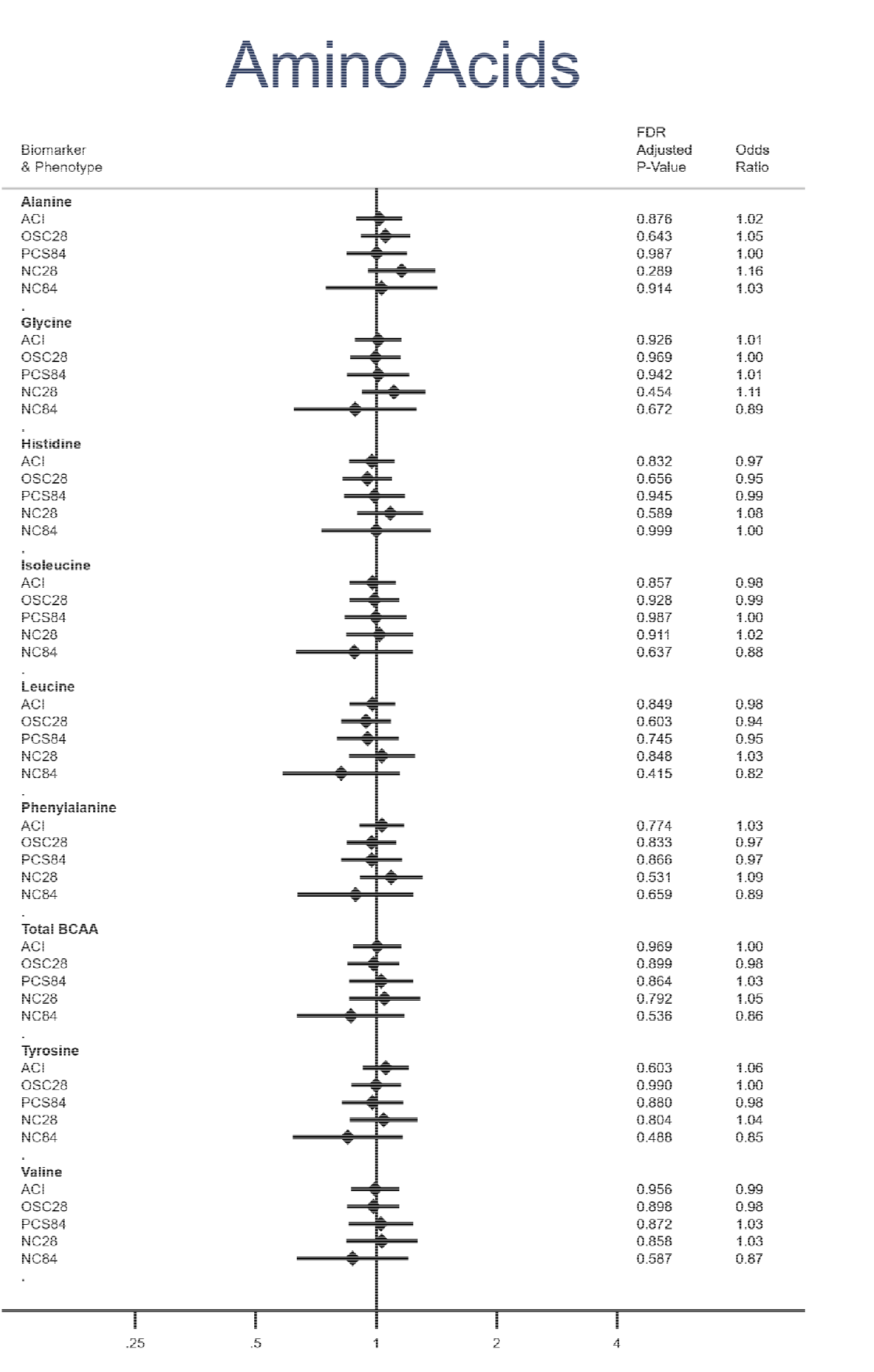
Relative risk ratio for each illness phenotype, per 1-SD increase in biomarker. Red indicates p≤0.05 after FDR correction Reference group (OR 1.0): Asymptomatic ACI: Acute COVID-19 illness OSC28: Ongoing symptomatic COVID-19 (28-83 days) PCS84: Post COVID-19 syndrome (≥84 days) NC28: Non-COVID-19 illness 28-83 days NC84: Non-COVID-19 illness ≥84 days LDL-C: Low density lipoprotein cholesterol HDL-C: High density Lipoprotein cholesterol Total-C: Total cholesterol Total-Tg: Total Triglycerides VLDL-C: Very low desnity lipoprotein cholesterol ApoA1: Apolipoprotein A1 ApoB: Apolipoprotein B

Amongst the subset of metabolites validated for clinical use (37 of 249 measurements)^18^, fatty acids differed in asymptomatic cases compared to both positive and negative symptomatic individuals. A higher ratio of polyunsaturated fatty acids (PUFA) compared to monounsaturated fatty acids (MUFA) was associated with a lower odds of prolonged COVID-19 (OR= 0.73 FDR-P=0.01 for OSC28 vs. asymptomatic), and non-COVID-19 illness (OR=0.68, FDR-P=0.01 for NC28 vs. asymptomatic). (Figure 1 and Supplementary Table 5). We also noted an association of absolute MUFA levels with increased length of illness (OR=1.28 [FDR-P=0.04] for OSC28 vs. asymptomatic COVID-19). Similarly, in combination, raised triglycerides and VLDL lipids were associated with an increased risk of prolonged illness in both test-positive and test-negative individuals, as was the ratio of triglycerides to phosphoglycerides (Supplementary Table 4 + 5). In contrast, higher HDL lipoprotein levels and larger HDL particles were associated with Asymptomatic cases. Neither amino acids nor glycoprotein acetyls were associated with COVID symptom duration.

In both the clinically validated variables, and the entire metabolomics dataset, only 7/249 (2.8%) variables were significantly different in Long COVID (OSC28 or PCS84 combined, Supplementary Table 6) compared to Non-COVID illness (NC28 and NC84 combined) as the reference group. These 6 of these 7 metabolites were not altered in asymptomatic and short COVID-19, and so may reflect differences specific to prolonged COVID illness. However, HDL-Cholesterol was also raised in Acute COVID-19 Illness (OR 1.24 [FDR P-value 0.005] for ACI vs Non-COVID illness) and further raised in Asymptomatic illness (OR 1.44 [FDR P-value 0.007] for Asymptomatic vs. Non-COVID illness).

### The Infectious diseases score

The multi-biomarker infectious diseases score (ID Score) was calculated, whereby higher values were previously associated with hospitalization for COVID-19^18^. In our cohort, higher values of the ID Score were generally associated with longer duration of symptoms of all illnesses, but notably not Post-COVID Syndrome (Table 3).

**Table 3:** Relative risk-ratios (RRR) for a 1 SD increased in ID Score, for each illness phenotype compared to asymptomatic COVID-19. Groups in bold are those for primary analysis.

| Group | RRR for ID Score | P-Value | 95% CI |
| --- | --- | --- | --- |
| **Asymptomatic** | **1.00** | (ref) | (ref) |
| **Acute COVID-19 Illness ($\leq 7$ days)** | **1.37** | **0.0032** | **(1.11 -1.68)** |
| Acute Non-COVID-19 Illness ($\leq 7$ days) | 1.31 | 0.0377 | (1.02 -1.68) |
| Intermediate COVID (8-27 days) | 1.35 | 0.0143 | (1.06 -1.72) |
| Negative Intermediate Illness (8-27 days) | 1.48 | 0.0013 | (1.17 -1.88) |
| **Ongoing Symptomatic Covid (28-83 days)** | **1.52** | **0.0002** | **(1.22 -1.90)** |
| **Non-COVID-19 Illness (28-83 days)** | **1.53** | **0.0022** | **(1.16 -2.00)** |
| **Post COVID-19 Syndrome ($\geq 84$ days)** | **1.24** | **0.1508** | **(0.92 -1.67)** |
| **Non-COVID-19 illness ($\geq 84$ days)** | **2.12** | **0.0006** | **(1.38 -3.27)** |

### Sensitivity Analyses adjusting for additional variables

The direction of effect and significance of results were unchanged for most sensitivity analyses; however, the effect of the ID Score was marginally stronger after adjusting for Healthy Plant-based diet index (OR=1.61 with hPDI, OR=1.53 without hPDI: for OSC28 compared to asymptomatic COVID-19) (Supplementary Figure 1).

### Microbiome demographics

A subset (n=301) of the metabolomic cohort had corresponding microbiome data (Table 2b). The median time between symptom onset and microbiome assessment was 223 (IQR 50 days), with the minimum time between symptom onset and microbiome assessment of 33 days (implications considered further in Discussion).

Quality control of gut microbiome sequence data for these 301 individuals revealed good breadth of coverage of MetaPhlAn markers. In addition, depth of coverage of MetaPhlAn markers was high for most abundant species (∼3X), with large areas of <0.5X coverage (Supplementary Figure 2).

For this subset analysis, individuals were grouped into categories as follows: 1) Asymptomatic: n=35; 2) ACI: n=109; 3a) OSC28 n=52; 3b) Post COVID-19 syndrome (≥84 days): n=20; 4) Negative symptomatic (≥28 days, including ≥ 84 days): n=85.

### Alpha-diversity analysis

Microbial richness did not differ between groups (Wilcoxon signed-rank test p-value>0.25 for all comparisons, Figure 2). There were no differences in Alpha-diversity (Simpson or Shannon) between groups, whether they were test positive or negative, symptomatic or asymptomatic, or with long or short symptom duration (Figure 2).

**Figure 2:**
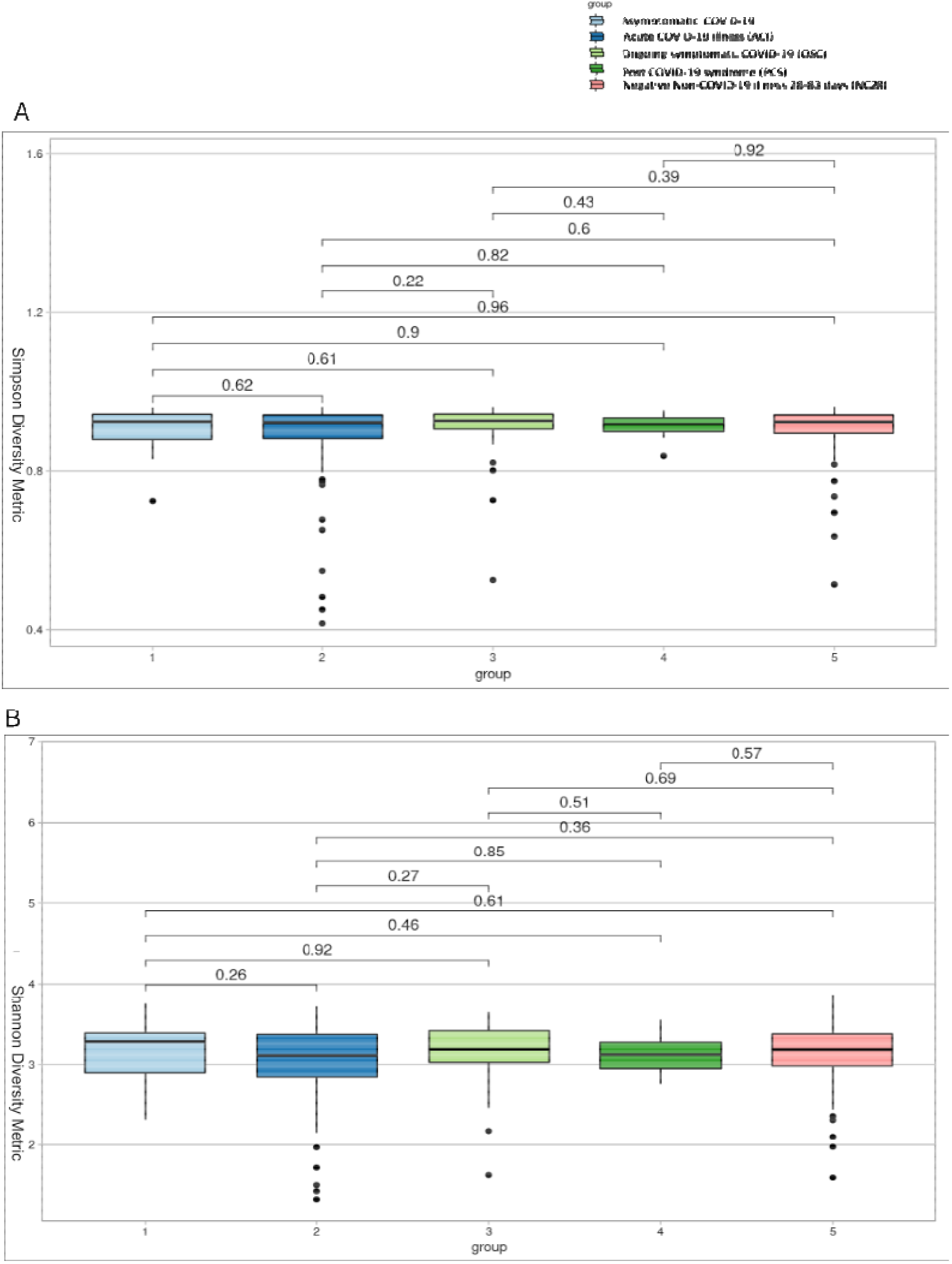
A) Simpson alpha-diversity and B) Shannon alpha-diversity. Simpson and Shannon alpha-diversity analysis showed no significant differences between groups. Species richness (Y-axis) was relatively high in COVID groups (X-axis). Wilcoxon signed-rank test with Dunn’s post hoc correction for multiple testing P-values are shown.

### Beta-diversity analysis

Similarly, beta-diversity analysis showed no large-scale shifts in microbial profiles between groups (Figure 3).Ongoing symptomatic COVID-19 and Post COVID-19 syndrome groups were amalgamated and beta-diversity analysis repeated; there remained no significant difference (data not shown).

**Figure 3:**
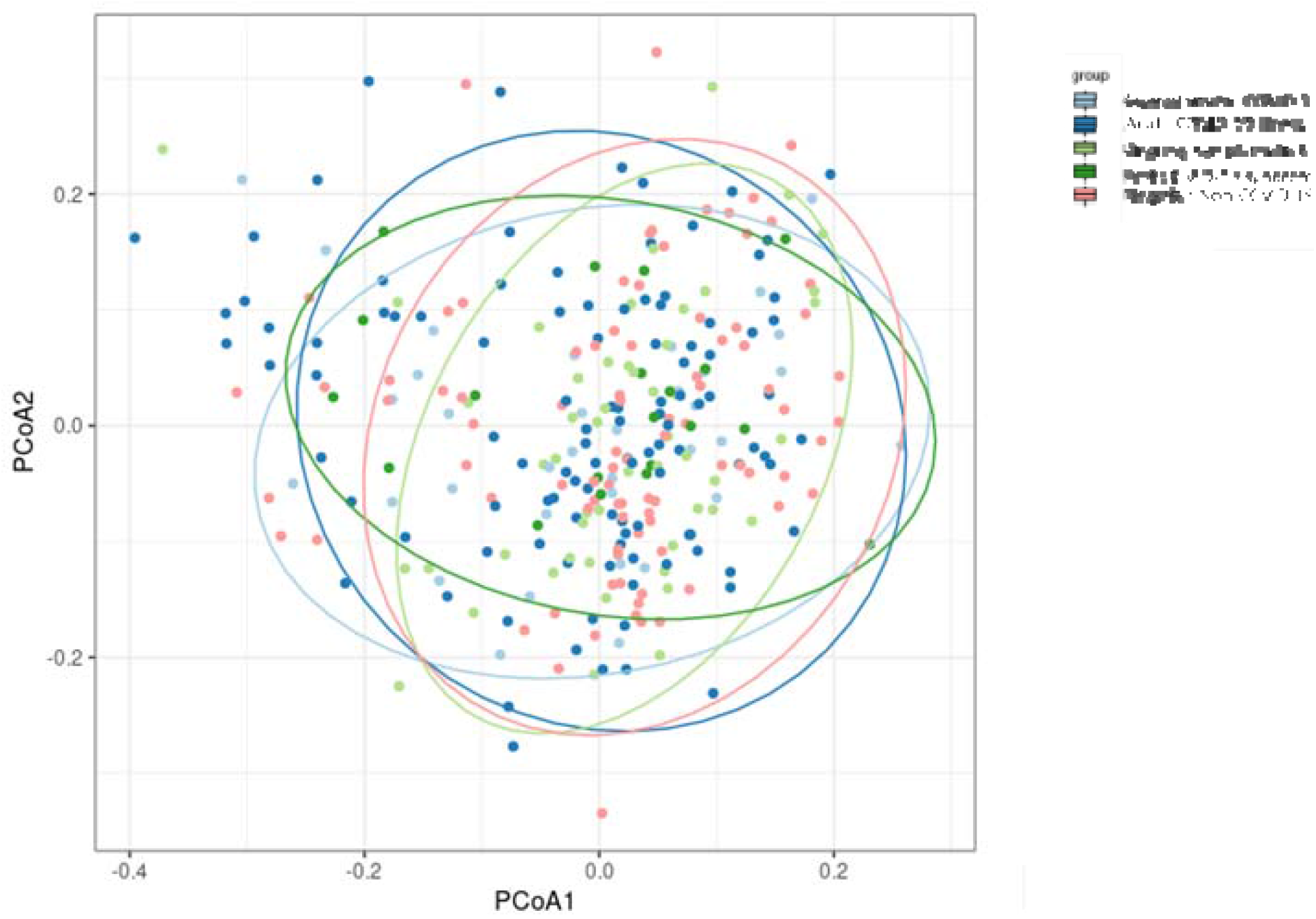
Principle Coordinate Analysis (PCoA) based on Bray Curtis dissimilarity metrics, showing the dissimilarity between five groups. among groups. Elipses represent 95% Confidence Intervals. Principal coordinate analysis (PCoA) ordination based on Bray-Curtis dissimilarity was used to visualize the dispersion of microbial community among groups. Elipses represent 95% Confidence Intervals.

### Biomarkers Asymptomatic

Kruskal-Wallis and Dunn’s post-hoc test identified three species with differences between groups - specifically, Firmicutes bacterium CAG 94, Ruminococcus callidus and Streptococcus vestibularis. Firmicutes bacterium CAG 94 differed between Asymptomatic SARS-CoV-2 infection (ref) and Acute COVID-19 (FDR P-Value = 0.03), OSC28 (FDR P-Value = 0.01) and PCS84 (FDR P-Value = 0.04). Ruminococcus callidus, also a Firmicute, differed between Asymptomatic and Non-COVID-19 illness (≥28 days) (FDR P-Value = 0.01) (See Table 4).

**Table 4:** Potential biomarkers most divergent between COVID groups. FDR corrected p-values <0.1 were considered significant. OSC28: Ongoing symptomatic COVID-19; PCS84: Post-COVID-19 syndrome; NC: Non-COVID illness _≥_28 days (including 28-83 days and _≥_84 days)

| Group Comparison | Z | P-Value | FDR P-Value |
| --- | --- | --- | --- |
| ***Streptococcus vestibularis*** |  |  |  |
| Asymptomatic - PCS84 | 2.115 | 0.034 | 0.114 |
| OSC28 - NC | -2.439 | 0.014 | **0.073** |
| PCS84 - NC | -2.549 | 0.010 | 0.107 |
| ***Ruminococcus callidus*** |  |  |  |
| Asymptomatic - ACI | 2.557 | 0.011 | **0.052** |
| Asymptomatic - PCS84 | 1.991 | 0.046 | 0.154 |
| Asymptomatic - NC | 3.254 | 0.001 | **0.011** |
| ***Firmicutes bacterium_CAG_94*** |  |  |  |
| Asymptomatic - ACI | 2.712 | 0.006 | **0.033** |
| Asymptomatic - OSC28 | 3.186 | 0.001 | **0.014** |
| Asymptomatic - PCS84 | 2.498 | 0.012 | **0.041** |
| Asymptomatic - NC | 2.058 | 0.039 | **0.098** |

### Correlation Between Metabolomic and Microbiome Analysis

Correlations between the metabolites and microbial taxa were assessed using Spearman’s rank coefficient. We found no evidence that microbiome taxa were associated with differences in those metabolites associated with symptom duration in this dataset (See Figure 4).

**Figure 4:**
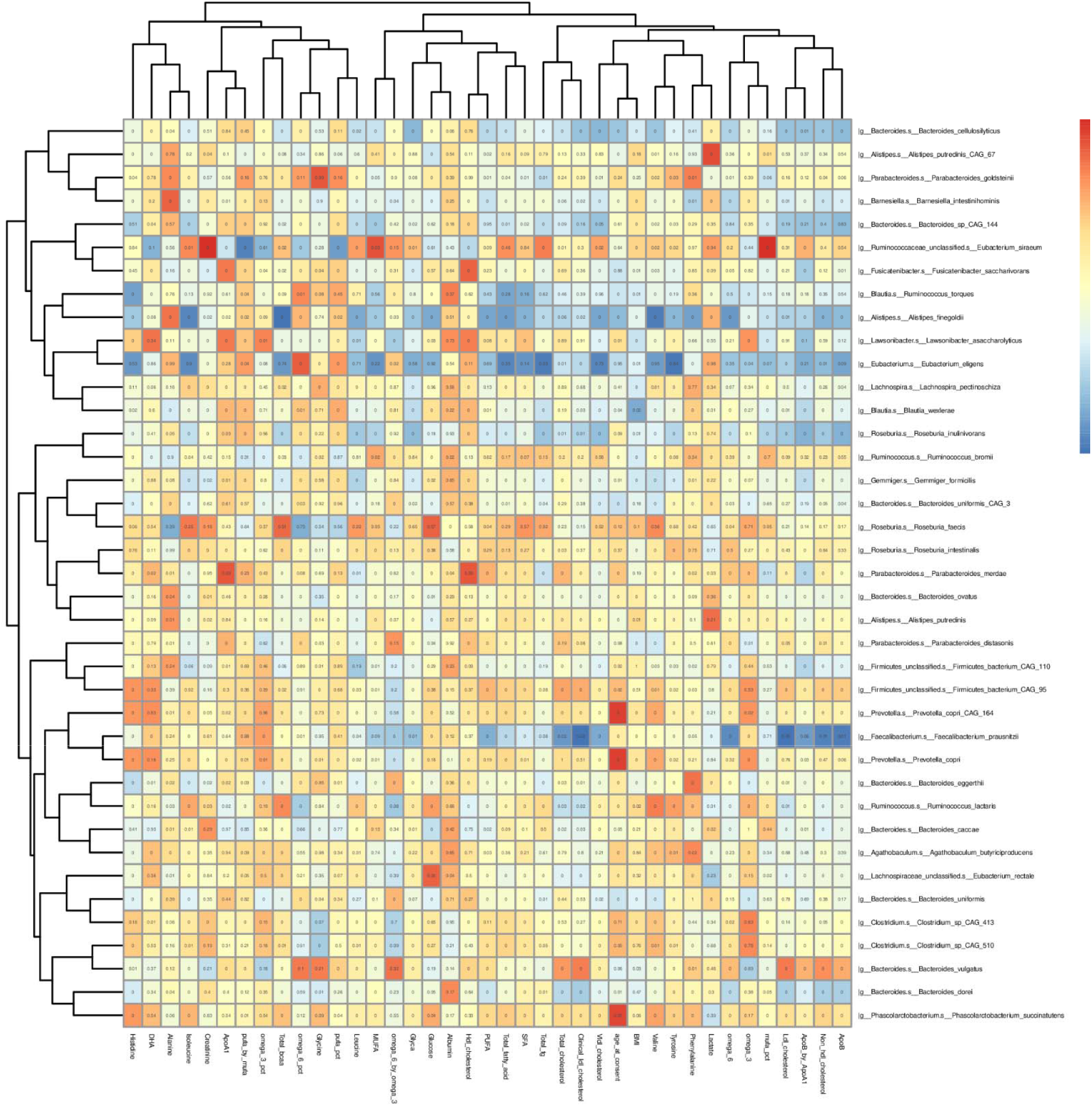
Spearman correlation of microbiome profiles and metabolomic profiles. Single microbial taxa correlated with clinically validated metabolomic data using Spearman’s rank sum non-parametric test. FDR p-values are displayed. Rows and columns are hierarchically clustered (Euclidean distance).

## Discussion

### Summary of Results + Results in Context

We observed a metabolic profile, particularly in lipid components, that differentiated individuals with longer symptom duration compared with individuals with asymptomatic infection. This profile was evident for individuals with long symptom duration regardless of SARS-CoV-2 test status, compared with asymptomatic infected individuals.

The specific differences identified in association with long-duration illness were small in magnitude and related to atherogenic dyslipidaemia (Supplementary Figure 3); in particular, blood fatty acid concentrations, including higher absolute and relative concentrations of MUFA, lower relative concentrations of PUFA, and a lower PUFA/MUFA ratios. In humans, circulating PUFA is derived from dietary sources, and blood levels correlate both to dietary intake and to levels in adipose tissue stores. MUFA, on the other hand, is synthesised in significant quantities in vivo and circulating concentrations (but not dietary intakes) are associated with increased risk of cardiometabolic disease^48^. Elevated serum MUFA and low serum PUFA have been identified in many studies as associated with ill health, including cardiovascular risk^49^, metabolic syndrome^50^, and mortality from infections^51^. Although our study assayed blood levels up to 9 months after initial COVID illness, these measures, particularly MUFA, are relatively stable over time^18,52^ and may reflect long-term blood concentrations. Moreover, our results concord with previous work in the UK Biobank, using the same platform, which demonstrated a similar direction of association with the same metabolites with increased risk of acute severe COVID-19 and with pneumonia, using blood samples collected many years beforehand^18^. Recent studies have reminded clinicians of the increased risk of vascular diagnoses after both COVID-19 and similar respiratory infections^53–55^ and while the risk reduces dramatically after the acute period, there is excess risk which remains many months afterwards. It is possible that our findings may reflect un-detected prior cardiovascular risk in symptomatic COVID-19 cases or vascular changes as a consequence of disease.

Higher levels of VLDL-particles/lipids and TG-enriched lipoproteins were associated with longer illness duration, although again effects were relatively small in absolute terms. Both components have also been associated with ill-health in other (pre-pandemic) studies - in particular, cardiovascular disease, diabetes, renal disease, obesity and depression^27,56^, although studies did not always adjust for diet^57^. In our study, adjusting for dietary intakes or co-morbidities did not change the findings. We also demonstrate similar findings to previous work associating peripheral vascular disease with metabolic profiles representing atherogenic dyslipidaemia (Supplementary Figure 3)^15,58^.

We did not find metabolomic profiles specific to Long COVID comparing individuals with long-duration illness who were positive vs. negative for SARS CoV-2 infection. Thus, our findings do not support a specific underlying metabolic alteration underlying long COVID per se; rather our work suggests that metabolic alteration is a risk factor for long symptom duration, regardless of the cause of illness. This is supported by a growing body of literature identifying “conventional” risk factors such as high BMI, diabetes and cardiovascular disease, as risk factors for long COVID, as well as age^7,59^ all of which are associated with adverse metabolic changes^49,56,60^. There was no meaningful change in our results with inclusion of self-reported cardiovascular disease and diabetes as co-variates (Supplementary Figure 1).

There were some unexpected negative findings. Others have associated GlycA with increased COVID-19 mortality^61^, and it is often increased in other conditions such as diabetes^56^. It was the biomarker showing the greatest association with hospitalisation for COVID-19 using pre-pandemic UK Biobank samples^18^. However, we found no association of GlycA with illness duration in our community-based sample. GlycA is considered a marker of inflammation, and our participants were sampled many months after the onset of symptoms and SARS-CoV-2, and were no longer reporting symptoms. It is possible that prolonged symptoms after COVID-19 may not always be related to ongoing inflammation, but previous damage that has not been repaired. There was also no association between any of the amino acid metabolites and length of illness, although some amino acids have previously been associated with hospitalisation for COVID-19^18^ and were included in the ID Score.

Looking at the ID score, previously associated with hospitalisation for acute pneumonia and acute COVID-19 illness^18^, we also demonstrated an increased score was associated with increased length of illness. This was replicated for both COVID-19 and those reporting ongoing symptoms without COVID-19. This suggests that there are shared associations which link severe acute illness and prolonged illness. This tallies with our previous observation that participants with a high acute symptom burden early in their illness were at greater risk of Long COVID^7^.

Although 97 metabolites and the ID score were associated with Ongoing Symptomatic COVID-19 illness, only 16 of these were still associated with Post-COVID-19 syndrome. The PCS84 group is much smaller, and may be heterogeneous. Further work is needed to see whether certain subgroups within PCS84 display similar associations to OSC28, whilst other subgroups do not. However, it is of note that those most at risk for PCS84, women in mid-life^4,62^, are less at risk for some conditions associated with metabolic derangement, such as cardiovascular disease^63,64^. Our results might suggest that truly persistent symptoms in PCS84 represent a different type of disease to OSC28, NC28 and NC84, with different risk factors, pathophysiology, and may therefore need different interventions to prevent or treat.

There was little change in the effect of the ID score on length of illness in our sensitivity analyses including pre-morbid cardiovascular disease and diabetes, frailty, lifestyle, IMD, the use of any supplements and the Diet Quality Score, suggesting that our metabolite associations with length of illness are independent of their associations with these variables. Models controlling for whether individuals took omega-3 supplements showed a diminished relationship between ID score and length of illness in non-COVID disease but were unchanged in COVID-19 illness. This finding which might suggest that omega-3 supplementation is a marker explaining this relationship in non-COVID illness only. The addition of the healthy plant-based diet index to the model increased the effect size of the ID score for each length of illness. Further work on hPDI and other correlated variables, such as socio-economic status, might help explore this finding.

We found no large-scale shift in gut microbe profile, assessed 9 months post symptom onset, in relation to illness duration after SARS CoV-2 infection, and notably, no difference between COVID-19 positive and negative individuals with long-duration symptoms. Analysis of both taxonomic and functional microbial profiles showed increases in relative abundance of some Firmicutes in asymptomatic individuals, for example Firmicutes bacterium CAG:94, an uncharacterized taxon requiring further study to determine its functions. This finding should be interpreted with caution, and we did not find evidence that it related to metabolite alterations. Here, timing of our sampling is highly relevant to the interpretation of our results. Alterations in an individual’s microbiome may occur in the context of acute disease,^65^ whether from infection, dietary changes, medication, usage, and/or immune function; however, an individual’s microbiome regresses over time ^66^ to reflect a stable baseline microbiome^67,68^. Thus, faecal microbiome samples collected 9 months after the start of illness may represent baseline individual microbiome, which, in our study, did not relate to symptom duration. In any case, our findings would suggest no long-term alterations in microbial profile in individuals who experienced ‘long COVID’. Our results contrast to one small previous study of 106 hospitalised individuals, of whom 76% had ongoing symptoms 6 months after acute SARS-CoV-2 infection. In this study, altered gut microbiome composition was reported in individuals with persistent symptoms in patients with COVID-19 compared to healthy historical pre-pandemic controls ^69^. However, cases were hospitalised for an average of 17 days, received amoxicillin-clavulanic acid among other interventions (including Ribavirin, Interferon and Remdesivir), and changes seen could have been a consequence these treatments, illness severity and/or altered diet in these individuals. Two other studies noted changes in immune-modulating commensal bacteria^70,71^ potentially specific to COVID-19, but again in hospitalised cohorts, where additional treatments, and illness severity, differ from our community-dwelling participants.

### Strengths + Limitations

Our study benefits from large size, with a long duration of prospective symptom reporting, and availability of both metabolite and microbiome data on the same community-based participants. These platforms are well validated, including for clinical use of the metabolomics data. Our participants also reflect the spectrum of COVID cases in the community. As participants were recruited prior to vaccination in the wild-type era, this reduces complexity by variance attributable to virus variant, and type and timing of vaccination^72^. Infection status was reconfirmed at enrolment, ensuring accuracy of classification by gold standard methodology. During this period, there were also relatively few acute COVID-19 specific treatments available, and none routinely used in UK community-managed individuals allowing our study to reflect the natural history of COVID-19.

We have also been able to conduct sensitivity analyses including variables such as frailty, lifestyle, deprivation, and diet, often not extensively accounted for in metabolomics studies, in addition to more traditional medical co-morbidities. We also have analysed participants who report ongoing symptoms not attributable to SARS-CoV-2, and therefore able to test the specificity of our findings.

Limitations include the cross-sectional nature of the metabolites and microbiome assayed up to 9 months after illness onset. Time of sampling, for both the metabolomics and microbiome analysis is both a strength and a limitation. Post convalescence sampling means that acute perturbations are likely to have resolved, but without pre-pandemic sampling we cannot assess whether changes found were consequential or pre-existing. Longitudinal data from cohorts sampled before and after pandemic are needed to address this issue.

## Conclusion

Metabolic profiles of community cases with asymptomatic COVID-19 were notably different to those with longer illnesses, displaying an atherogenic lipoprotein phenotype, and this difference was apparent regardless of whether the illness was due to COVID-19 or another acute phenomenon. Those with COVID-19 symptoms for ≥28 days could not be clearly distinguished from those with non-COVID-19 illnesses of prolonged duration. A biomarker score previously predictive of severe COVID-19 was overall predictive of prolonged illness, although not all individual components were. In contrast, gut microbiome diversity did not differ by length of illness, suggesting no significant gut microbiome dysbiosis post COVID-19 infection.

Further research with longitudinal sampling pre- and post-illness is warranted, to determine if the observed metabolomic associations with longer illness are pre-existing risk factors, or consequential.

## Data Availability

Data collected in the ZCS smartphone application are shared with other health researchers through the UK National Health Service-funded Health Data Research UK (HDRUK) and Secure Anonymised Information Linkage consortium, housed in the UK Secure Research Platform (Swansea, UK). Anonymised data are available to be shared with researchers according to their protocols in the public interest (https://web.www.healthdatagateway.org/dataset/fddcb382-3051-4394-8436-b92295f14259). The code is available in: https://gitlab.com/KCL-BMEIS/covid-zoe/vaccination. Access to data in the CSS Biobank is available to bona fide health researchers on application to the CSS Biobank Management Group. Further details are available online at: https://cssbiobank.com/information-for-researchers including application forms and contact information.

https://gitlab.com/KCL-BMEIS/covid-zoe/vaccination

https://web.www.healthdatagateway.org/dataset/fddcb382-3051-4394-8436-b92295f14259

## Acknowledgments

We would like to acknowledge support from the Exetera development team (Benjamin Murray, Liyuan Chen, Jie Deng, Eric Kerfoot), the TwinsUK laboratory and administrative teams (Gulsah Akdag, Andy Anastasiou, Andrei Beleanu, Julia Brown, Maria Paz Garcia, Rachel Horsfall, Genevieve Lachance, Ayrun Nessa, Alyce Sheedy, Dovile Vaitkute, Sam Wadge, Sivasubramaniam Rajan Wignarajah, Darioush Yarand). We are also grateful to the CSS Biobank Volunteer Advisory Group for their input on the development of the biobank, and feedback on the significance of this work for affected individuals and the wider public.

