## Supplementary Methods and Figures for "Metabolomic and gut microbiome profiles across the spectrum of community-based COVID and non-COVID disease: A COVID-19 Biobank study"

#### DNA Extraction

Briefly, 1g faecal sample was mixed with 5-6 ml of distilled water and 2 Core 5 mm glass beads. Tubes were put in Spex Grinder for 10 sec at 800. Contents settled for 12 mins, supernatant transferred to Core beat tube and centrifuged for 10 min at 15,000 g. Supernatant was removed and 400µl Core Clarifying solution was added. Bead beating of samples in Spex Grinder for 5 minutes at a rate of 1000. Samples were centrifuged for 3 minutes at 15,000 g. Proteinase K (10µl) was added to the deep well plate. 200µl of each sample was added to each well and mixed thoroughly. Finally, 720µl of Lysis/Bind Master Mix was added to each well. Plates were then run using King-Fisher Flex, as per manufacturer’s instructions. Elution was in 100µl of MagMax Core elution buffer. gDNA quantity was estimated using Qubit.

### Supplementary Figures

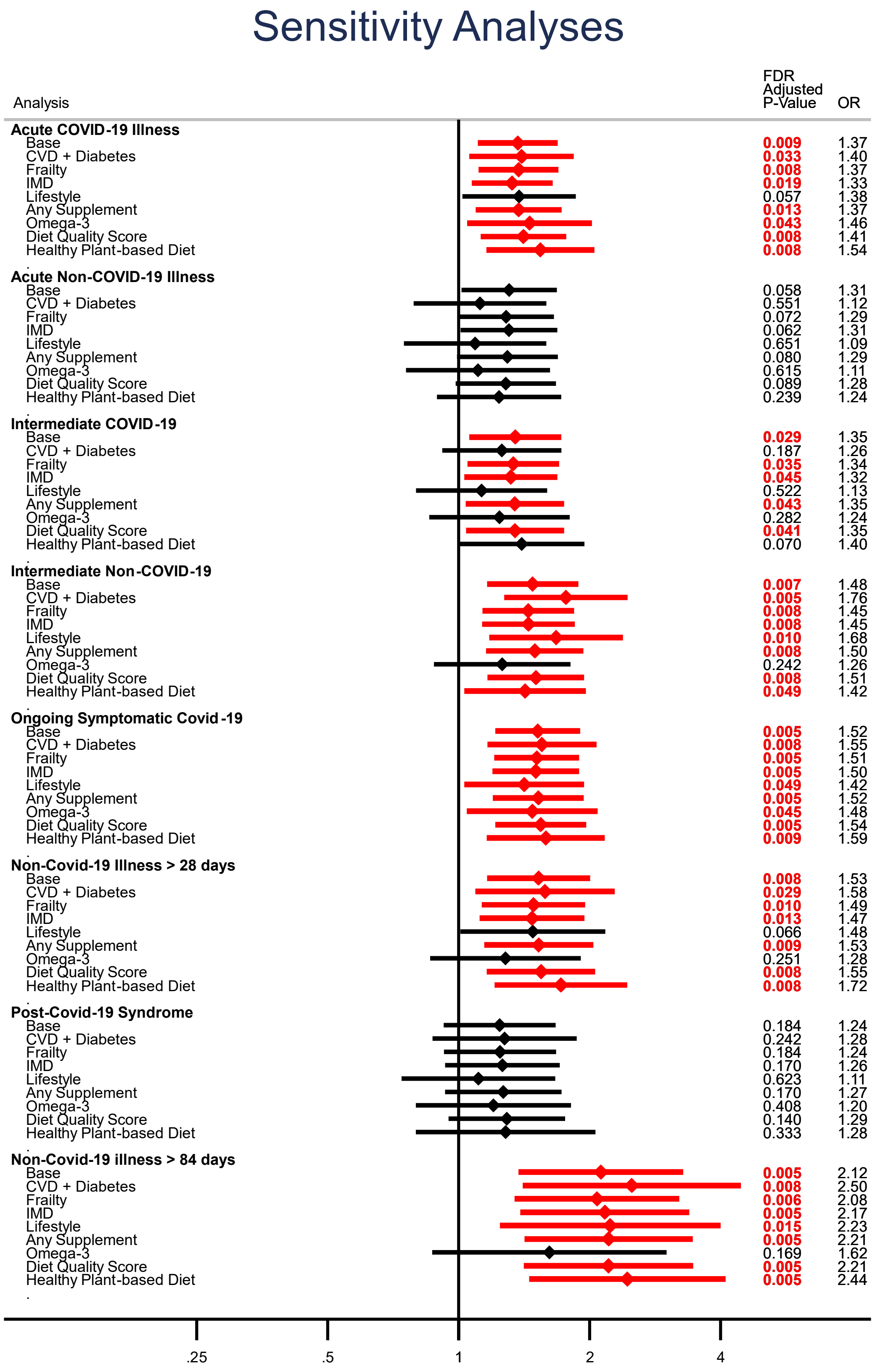


**Supplementary Figure 1:** Sensitivity analyses comparing the effect of additional variables on the risk of illness phenotype, compared to asymptomatic Covid-19.

Base is our model adjusted for age, sex and body mass index.

95% Confidence intervals displayed with p-values adjusted using Benajmini-Hochberg False Discovery Rate correction.

Red indicates FDR corrected p-value <=0.05

CVD – Self-reported cardiovascular/Heart disease;

IMD – Index of Multiple Deprivation;

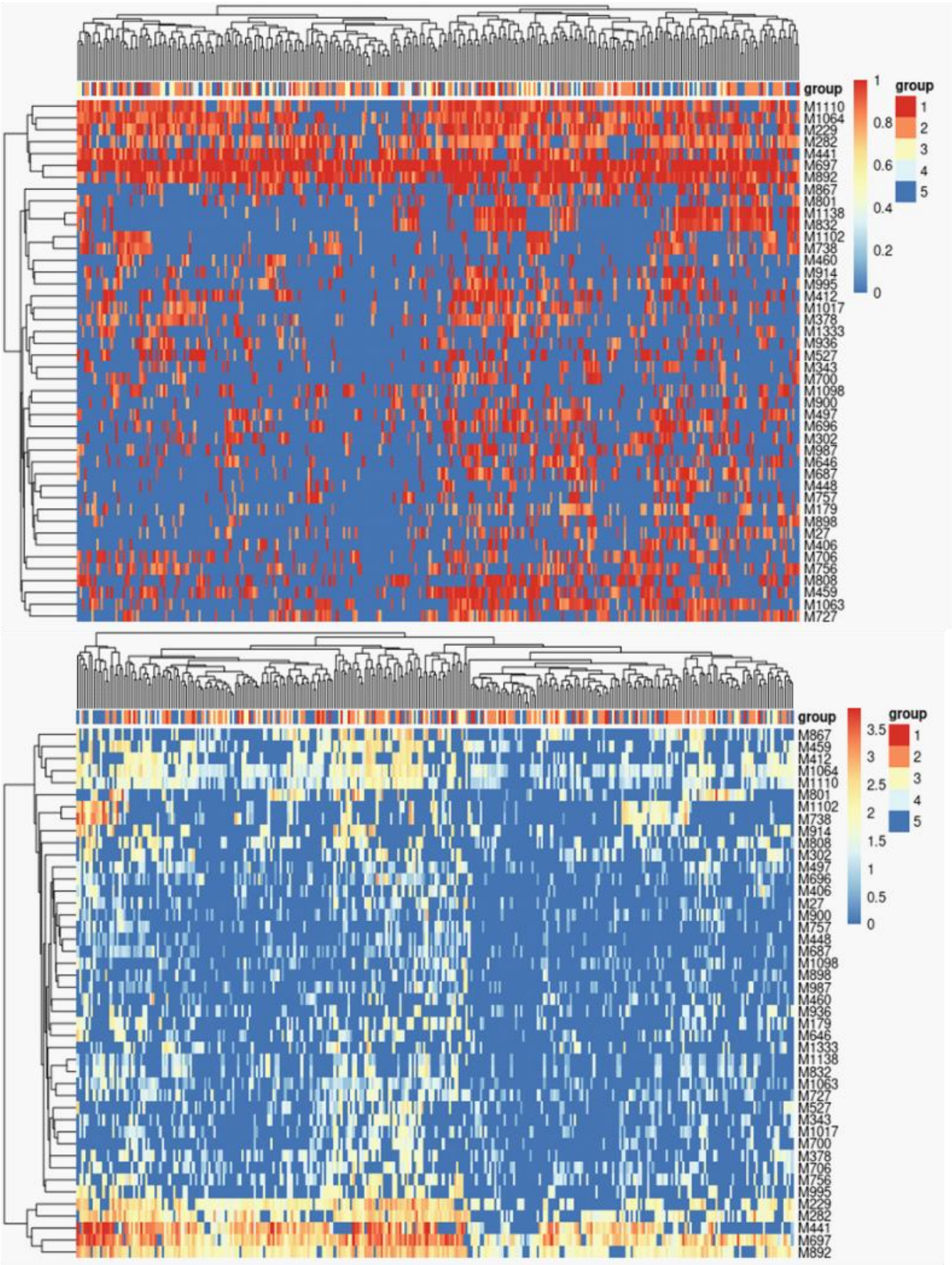


**Supplementary Figure 2:** Quality control of A) Breadth of coverage and B) Depth of Coverage for microbiome sequencing cohort. Sample (x-axis) and markers (Y-axis) are coloured based on breadth of coverage (ensures marker genes are sequenced) and depth of coverage (determines depth of sequencing).

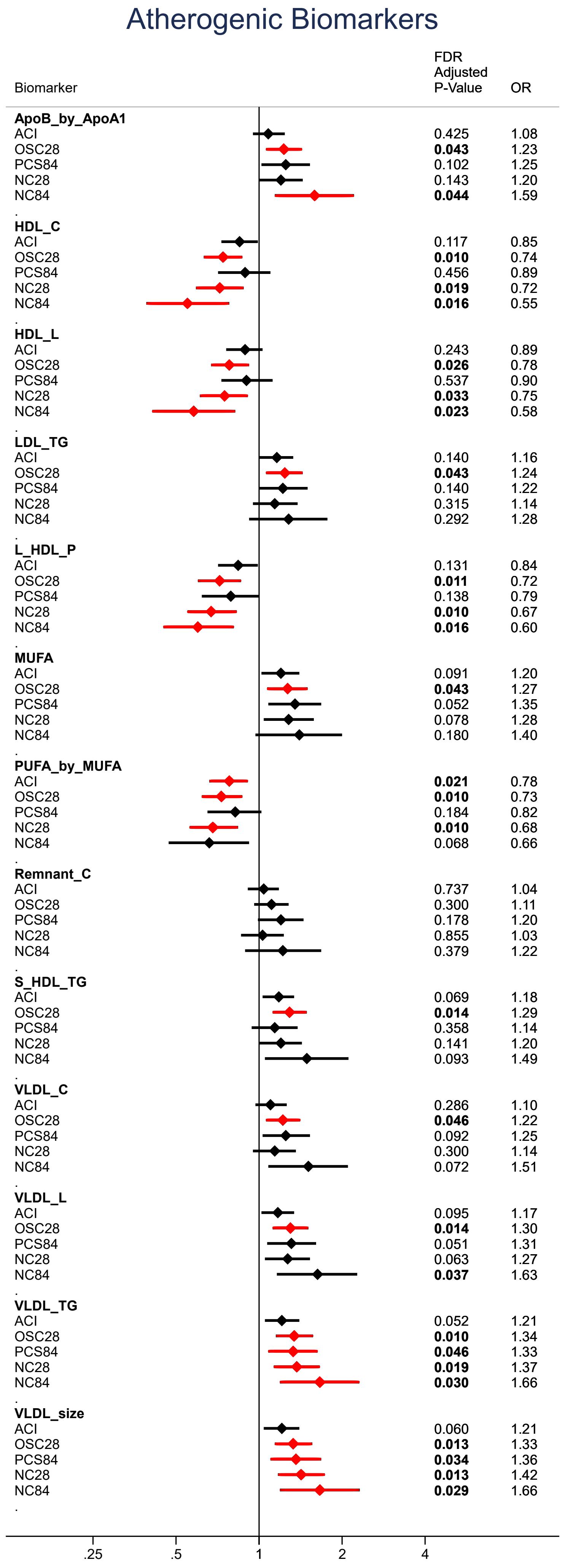


**Supplementary Figure 3:** Atherogenic-dyslipidaemic biomarkers.
Relative risk ratio for each illness phenotype, per 1-SD increase in biomarker.
Adjusted for age, sex and body mass index.
95% Confidence intervals displayed with p-values adjusted using Benajmini-Hochberg False Discovery Rate correction.

Red indicates FDR corrected p-value <=0.05

ACI: Acute COVID-19 illness
OSC28: Ongoing symptomatic COVID-19 (28-83 days)
PCS84: Post COVID-19 syndrome (≥84 days)
NC28: Non-COVID-19 illness 28-83 days
NC84: Non-COVID-19 illness ≥84 days

ApoB_by_ApoA1: Ratio of apolipoprotein B to apolipoprotein A1
HDL_C: High density lipoprotein cholesterol
HDL_L: Total Lipids in high density lipoprotein
LDL_TG: Triglycerides in low density Lipoprotein
L_HDL_P: Concentration of large high density lipoprotein particles
MUFA: Monounsaturated Fatty Acids
PUFA_by_MUFA: Ratio of polyunsaturated fatty acids to monounsaturated fatty acids
Remnant_C: Remnant cholesterol (non-HDL, non-LDL -cholesterol)
S_HDL_TG: Cholesterol in small HDL
VLDL_C: Very low density lipoprotein cholesterol
VLDL_L: Total lipids in VLDL
VLDL_TG: Triglycerides in VLDL
VLDL_size: Average diameter for VLDL particles
